# Factors associated with severe sepsis in diarrheal adults and their outcome at an urban hospital, Bangladesh: A retrospective analysis

**DOI:** 10.1101/2021.03.03.21252843

**Authors:** Monira Sarmin, Monjory Begum, Farhana Islam, Farzana Afroze, Lubaba Shahrin, Sharifuz zaman, Tahmina Alam, Abu Sadat Mohammad Sayeem Bin Shahid, Tahmeed Ahmed, Mohammod Jobayer Chisti

## Abstract

**Background:** Clinical features of sepsis and severe diarrhea often overlap and create a dilemma among the clinicians. To describe factors associated with severe sepsis in diarrheal adults and their outcomes to understand their interplay as clinical features of sepsis and severe diarrhea often overlap.

**Methods and results:** We used this retrospective chart analysis employing a case-control design to study critically ill diarrheal adults aged ≥ 18 years treated in ICU Dhaka hospital, icddr,b between January 2011 to December 2015. Diarrheal adults with a diagnosis of severe sepsis were cases and an equal number of randomly selected non-septic patients were the controls. Of 8,863 in-patient adults, 350 fulfilled the criteria of cases. Cases died significantly more (9% vs 3%, p=0.002) than controls. 69% of the cases progressed to septic shock. In logistic regression analysis, steroid intake, ileus, acute kidney injury, metabolic acidosis, and hypocalcemia were significantly associated with severe sepsis in diarrheal adults (all, p<0.05). 12% of cases (40/335) had bacteremia. *Streptococcus pneumoniae* [9 (22.5%)] was the single most common pathogen and gram-negatives [27 (67.5%)] were prevailing as a group.

**Conclusion:** Diarrheal adults who had ileus, AKI, metabolic acidosis, hypocalcemia, and also took steroids were prone to have severe sepsis. Strikingly, gram-negatives were the predominant bacteria among the diarrheal adults having severe sepsis.

## Introduction

Sepsis with its unpredicted cryptic course can affect any patient anywhere in the world. Sepsis results from a disrupted inflammatory response to infection in the body where it can progress to septic shock followed by multiple organ dysfunction syndrome and death [1]. Worldwide, the burden of sepsis is perceived to be high, though it remained understudied at 87% of the world population residing in lower-middle-income countries. A study including data from 1979 to 2015 projected a global burden of annual 31.5 million sepsis, and 19.4 million severe sepsis, and among them, 5.3 million died [2]. Studies from Asian critical care facility including Bangladesh reported poor compliance of the surviving sepsis campaign (SSC) bundles and the case-fatality-rate was as high as 49·2% from severe sepsis [3-4].

Dhaka hospital of icddr,b treats diarrheal adults, some of them also have severe sepsis and often experience fatal outcomes. Patients typically present with features of sepsis evident by infection, tachycardia, fever, and leukocytosis. Progression of sepsis resulting in hypotension and/or absent peripheral pulses from poor peripheral perfusion in the absence of dehydration is termed as severe sepsis. Unresponsiveness to isotonic fluid (30 ml/kg bolus of normal saline/ringer lactate over 10–15 min) and require the support of inotropes/vasopressors, is termed as septic shock. Oliguria, acute kidney injury and altered mental status signify the presence of organ dysfunction [5, 6]. following SSC recommendation, we administer the first antibiotic early that helps to reduce in-hospital mortality [7].

Thus, we need to predict severe sepsis at its early stage and prompt management of severe sepsis will reduce deaths in these adults by eliminating its progression to septic shock. However, data are lacking in diarrheal adults having sepsis, or consequences of sepsis in adults hospitalized for diarrhea. Thereby, we aimed to portray the clinical findings with the outcome of severe sepsis in diarrheal adults comparing them with non-septic adults.

## METHODS

### Study design

We retrospectively analyzed data of diarrheal patients aged ≥ 18 years and treated in icddr,b Dhaka hospital between January 2011 to December 2015, where cases had severe sepsis along with diarrheal illness and the controls had only diarrhea. Infection or the presumed presence of infection plus tachycardia plus hyperthermia (≥ 38.5°C) or hypothermia (≤35.0°C), or abnormal white blood cell number are the criteria of sepsis. A combination of sepsis and poor peripheral perfusion evident by hypotension and/or absent peripheral pulses without dehydration constituted severe sepsis [8]. Fluid (bolus of 30 ml/kg normal saline/ringer lactate over 10–15 min) unresponsiveness and requirement of inotropes confirmed the presence of septic shock. We compared the clinical characteristics of diarrheal adults with and without severe sepsis. We had 350 cases and an equal number of controls were chosen for comparison at random using a statistical package for the social sciences [IBM SPSS Statistics for Windows, Version 20.0. Armonk, NY: IBM Corp].

Every year, clinicians at Dhaka hospital manage approximately 150,000 patients having diarrhea, with or without other complications. Here almost every patient presented with diarrhea. We used their data for this chart analysis. A description of the Dhaka hospital has been illustrated in other papers [9].

ICU physicians re-evaluated patients having severe sepsis, started the required workup, and prescribed a standard management plan following the hospital’s guidelines. A portable pulse oximeter (OxiMaxN-600) measured capillary oxygen saturation (SpO_2_) and Accu Chek Active (Roche Diagnostics GmbH, Mannheim, Germany) glucometer estimated blood glucose. device. Hypoxemic patients received oxygen therapy (@4-5 liter/minute). In alignment with SSC 2012 guideline, hospital protocol has been developed. Before instituting antibiotics (within one hour of diagnosis) e.g., 3^rd^ generation cephalosporin and aminoglycosides (fluoroquinolone instead of aminoglycosides for concomitant community-acquired pneumonia), blood was drawn for culture and sensitivity and other investigations. Appropriate feeding (nothing by mouth with maintenance fluid for patients with septic shock), was provided as and when required. They received 30 ml/kg fluid bolus (normal saline/Hartmann) (25) and also received vasopressor and inotropes for septic shock evident by unresponsiveness to a fluid bolus. We started noradrenaline first, 0.05 microgram/kg.min and increased the dose after 15 minutes to 0.1 microgram/kg.min to a maximum of 0.5 microgram/kg.min. We used definitive response criteria: mean arterial pressure (MAP) ≥ 65 mm Hg, urine output (UO) ≥ 0.5mL/kg/hour, and central venous pressure (CVP) 8 to 12 mm Hg[10-11]. As we have limited expertise, we used to measure CVP in the external jugular vein, which is reliable, instead of the internal jugular vein. [12]. Thereafter, we added injection adrenaline if the goal was not achieved. In inotrope resistant shock, we also added injection hydrocortisone. Monitoring of the heart rate, respiratory rate, skin color/CRT, temperature, pulse oximetry, and mental status [13] are ensured besides the aforementioned criteria.

### Data management

We have collected the data from a computer-based record-keeping network, SHEBA. We provide a unique number, and against this number, all the data were recorded. We prepared the case report forms and finalized for data accession after pretesting. We collected demographics (age, gender), clinical information (presence of fever, cough, respiratory distress, disorientation, and their duration, history of [H/O] antibiotic use for the current illness, systemic steroid intake, and comorbidities e.g., chronic lung disease, diabetes mellitus, abnormal auscultatory findings in the lungs, and severe sepsis), and laboratory data (blood, and stool cultures and sensitivity, total white blood cell, serum creatinine, serum electrolytes, calcium and magnesium,) from the medical chart. We presented laboratory data as leukocytosis (white blood cell count >11,000/L) hyponatremia (serum sodium <135 mmol/L), hypokalemia (serum potassium <3.5 mmol/L), hypocalcemia (serum total calcium <2.12 mmol/L), hypomagnesemia (serum total magnesium <0.65 mmol/L), hypoglycemia (random blood sugar <3.0 mmol/L), metabolic acidosis (serum TCO_2_ <24 mmol/L) and acute kidney injury (AKI) if serum creatinine was 1.5 times the upper limit of normal, [Normal serum creatinine is 159 µmol/L in men and 146 µmol/L in women]. We also documented the hospital course and outcome and the number of days in the hospital.

### Data Analysis

We entered all the data into SPSS for Windows and Epi Info (version 7.0, Epi Info™software; Center for Disease Control and Prevention, Atlanta, GA, USA). We reported categorical data, like numbers and percentages; continuous data as means with standard deviations or medians with interquartile ranges (IQRs) as appropriate. The Student’s t-test compared means of homogenous data and the Mann-Whitney U test compared inhomogeneous data between the groups. Odds ratio (OR) and their 95% confidence intervals (CIs) were used to demonstrate the strength of association. For statistically significant, a p-value is set <0.05. To identify factors for severe sepsis in diarrheal adults, initially, a bivariate model was used, and then a multivariable logistic regression analysis model identified factors independently associated with severe sepsis after controlling for the relevant confounding variables.

### Ethical considerations

In this study, we reviewed only the medical records without involving any interviews with patients or caregivers. Data were anonymized before analysis. Nevertheless, the Institutional Review Board of icddr,b approved the study.

## Results

During the study period, out of 8,863 diarrheal inpatients at Dhaka hospital, icddr,b, 350 were the cases and thus, 350 were controls. Cases had significantly higher mortality than controls (Table1).

On admission, the cases had lower mean age, H/O systemic steroid intake before this illness, disorientation, ileus, pneumonia (Table 1), hypoglycemia, acute kidney injury (AKI), metabolic acidosis, hypokalemia, hypomagnesemia, and hypocalcemia (Table 2) and required long course in the hospital to recover (Table 1) compared to the controls. While in-patient, 240 (69%) cases developed septic shock and required inotropes (Table 1).

**Table 1.**
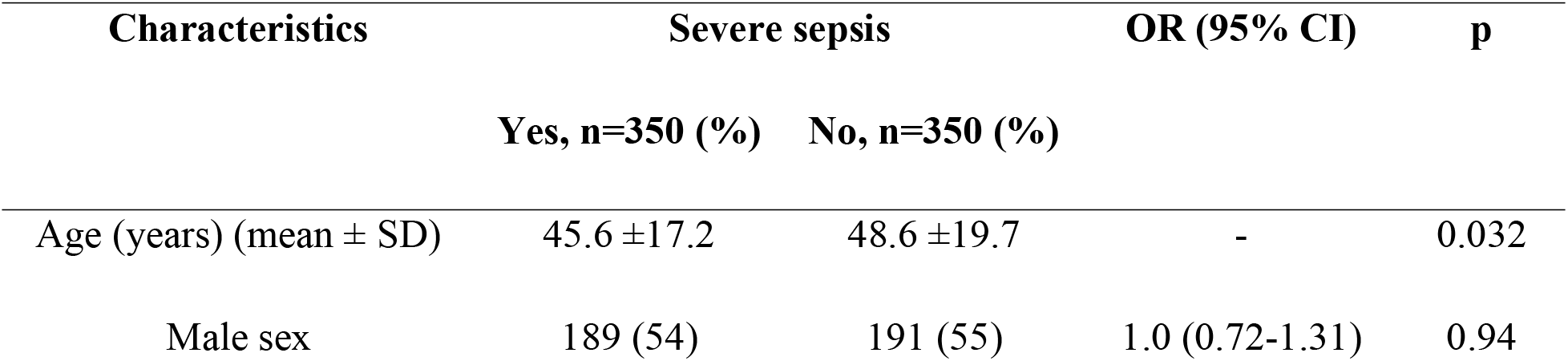
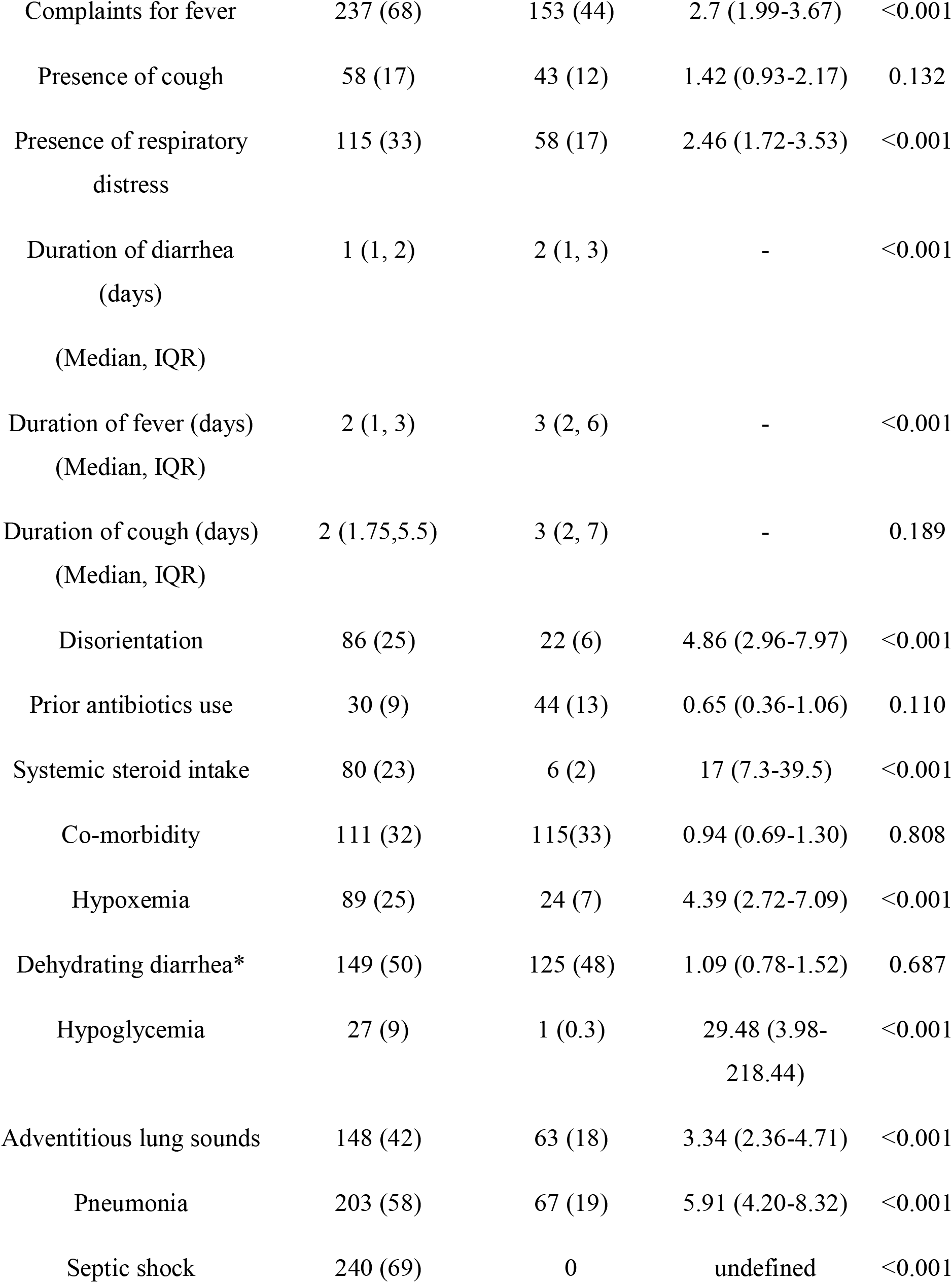
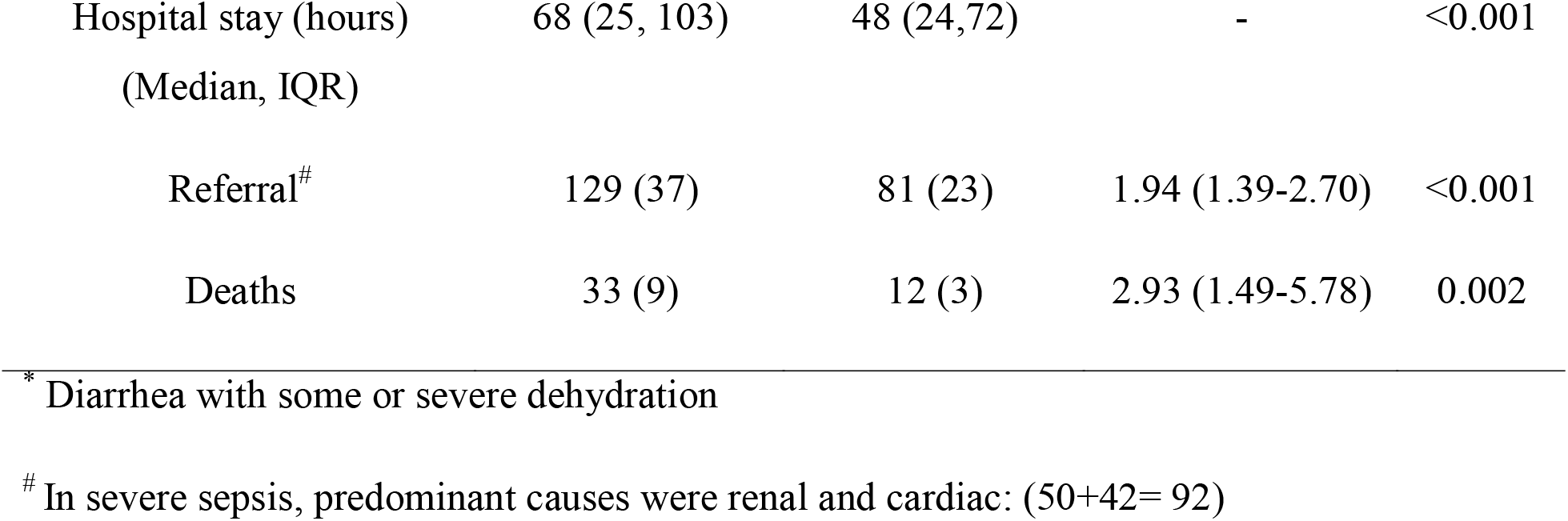
Clinical characteristics of diarrheal patients with and without severe sepsis.

**Table 2.** Laboratory characteristics of diarrheal patients with and without severe sepsis.

| Characteristics | Severe sepsis |  | OR (95% CI) | p |
| --- | --- | --- | --- | --- |
|  | Yes, n=350 (%) | No, n=350 (%) |  |  |
| Hb (gm/dl) (mean ± SD) | 12.13±2.59 | 12.3 ±2.74 | - | 0.344 |
| Hyponatremia | 94 (27) | 74 (30) | 0.87 (0.61-1.25) | 0.511 |
| Metabolic acidosis | 306 (90) | 171 (70) | 3.63 (2.33-5.64) | <0.001 |
| Hypokalemia | 94 (29) | 90 (38) | 0.66 (0.47-0.95) | 0.030 |
| AKI<sup>*</sup> | 223 (66) | 80 (34) | 3.75 (2.64-5.32) | <0.001 |
| Hypocalcemia | 274 (87) | 35 (55) | 5.34 (3.06-10.00) | <0.001 |
| Hypomagnesemia | 229 (74) | 29 (49) | 2.99 (1.69-5.31) | <0.001 |
| Invasive diarrhea | 120 (62) | 133 (64) | 0.94 (0.63-1.41) | 0.842 |
| Blood isolates | 40 (12) | 27 (18) | 0.63 (0.37-1.07) | 0.113 |
| Stool isolates | 22 (21) | 26 (23) | 0.89 (0.47-1.68) | 0.838 |

\*AKI=Acute kidney injury (if serum creatinine is 1.5 times the upper limit of normal)

The blood and stool isolate among the cases and the controls are shown in Tables 3 and 4 respectively. Among the cases, only 12% had bacteremia (40/335). As a single pathogen *Streptococcus pneumoniae* (22.5%) was the predominant bacterial isolates whereas as a group, gram negatives (67.5%) were prevailing (Table 3).

**Table 3.** Bacterial isolates from the blood of the study patients.

| Blood isolates | Severe sepsis |  |
| --- | --- | --- |
|  | Yes, n=40 (%) | No, n=27 (%) |
| *S. pneumoniae* * | 9 (22.5) | 0 |
| *S. Typhi* | 4 (10) | 14 (52) |
| *S. paratyphi* | 2 (5) | 0 |
| *Pseudomonas* spp | 7 (17.5) | 2 (7) |
| *E. coli* | 2 (5) | 5 (18) |
| *Enterobacter* spp | 6 (15) | 1 (4) |
| *Klebsiella* spp | 3 (7.5) | 0 |
| *Enterococcus* spp | 2 (5) | 1 (4) |
| *Acinetobacter* spp | 1 (2.5) | 3 (11) |
| *S. aureus* * | 4 (10) | 1 (4) |

\* *S. pneumoniae* and *S. aureus* are gram-positive (32.5%) and the rest are gram-negatives in the severe sepsis group

**Table 4.** Bacterial isolates from the stool of the study patients.

| Stool isolates | Severe sepsis |  |
| --- | --- | --- |
|  | Yes, n=22 (%) | No, n=26 (%) |
| *Shigella* spp | 8 (36) | 7 (27) |
| Salmonella group | 4 (18) | 8 (31) |
| campylobacter | 2 (9) | 3 (11.5) |
| *Vibrio* | 6 (27) | 5 (19) |
| *Aeromonas* | 2 (9) | 3 (11.5) |

In logistic regression analysis, systemic steroid intake, ileus, acute kidney injury (AKI), metabolic acidosis, and hypocalcemia were independently associated with severe sepsis (Table 5) where we adjusted the potential confounders.

**Table 5.** Logistic regression (Backward Conditional) analysis to explore the clinical predictors of septic shock.

| Characteristics | OR | CI | P |
| --- | --- | --- | --- |
| Systemic steroid intake | 8.87 | 1.55-30.41 | 0.011 |
| Disorientation | 1.20 | 0.53-2.72 | 0.664 |
| Hypoglycemia | 3.48 | 0.41-29.74 | 0.255 |
| Pneumonia | 1.55 | 0.74-3.25 | 0.241 |
| Ileus | 2.95 | 1.43-6.09 | 0.003 |
| Metabolic acidosis | 2.83 | 1.10-7.29 | 0.031 |
| AKI | 2.14 | 1.03-4.42 | 0.041 |
| Hypocalcemia | 8.20 | 3.79-17.75 | <0.001 |
| Hypokalemia | 0.53 | 0.26-1.08 | 0.080 |
| Hypomagnesemia | 1.56 | 0.70-3.52 | 0.279 |

OR=odds ratio, CI= confidence interval

## Discussion

We studied diarrheal adults to explore clinical and laboratory factors associated with severe sepsis and their outcome. We have also evaluated the spectrum and frequency of bacteremia in this population. Overall, the predominance of gram negatives among diarrheal adults having severe sepsis is the striking observation of this study is. Other important observations of our study were: (i) progression to septic shock from severe sepsis in diarrheal adult was high (69% [240/350]), (ii) Mortality rate was significantly higher among patients having severe sepsis and diarrhea compared to those without severe sepsis/sepsis, (iii) systemic steroid intake, ileus, acute kidney injury (AKI), metabolic acidosis, and hypocalcemia were independently associated with severe sepsis.

The predominance of gram negatives among the bacterial pathogens isolated from diarrheal adults having severe sepsis is remarkable. We may speculate that in diarrheal adults’ breaches of healthy intestinal flora might allow potential translocation of gram-negative pathogens to the bloodstream. Except for severe sepsis by *Streptococcus pneumoniae* from a respiratory source, gram negatives were found to be common pathogens in several previous studies involving adults with sepsis/severe sepsis and conducted in Southeast Asia and Europe [14-15].

Using the diagnostic criteria suggested by SSC guidelines, we found the proportion of septic shock among adults having severe sepsis was 69%, which is consistent with a recent study [16]. When diarrheal patients developed severe sepsis, their risk of death increases significantly. Among ICU patients, it is the 2^nd^ leading cause of death [15]. The mortality rate for sepsis in different studies varies and could be as high as 56% [15,17]. The deleterious cascade of sepsis causes multiorgan failure and death. Thus, the relationship between increased mortality and septic shock is well established [18,19]. Our observation of 9% mortality among diarrheal adults with severe sepsis might reflect good adherence to SSC guidelines. On the other hand, some patients required referral to different facilities for other associated illnesses as we did not have the facilities to treat those. Although we did not know their outcome, it is postulated that the mortality rate among them might also be higher.

We observed patients with an H/O systemic glucocorticoid intake had a significantly higher risk of severe sepsis when they also had diarrhea. Glucocorticoids are alluring drugs with their beneficial role for endocrine, inflammatory, allergic, and immunological disorders [20]. Besides, they increase the probability of infection with common bacteria, viruses, and fungus by impairing phagocyte function [21,22] and producing hypogammaglobulinemia even with short courses from outpatient settings [23]. Our observation of the association of ileus with severe sepsis is also understandable. Bacteria may translocate into the systemic circulation from an insulted intestine that may cause sepsis [24]. Diversely, during sepsis, splanchnic hypoperfusion and gut-derived mediators induced leukocytes malfunction hamper gastrointestinal motility and result in paralytic ileus [25].

We also observed a significant number of patients suffered from acute kidney injury (AKI). 66% of our study patients with severe sepsis developed acute kidney injury (AKI), which is consistent with previous observations [26-27]. Septic AKI is a consequence of the renal microvascular dysfunction, the interaction between pathogen fragments and renal cells, cytokine storm, or disintegration between injured organs [28]. However, after the restoration of the target mean arterial pressure (MAP), most of our study patients recovered from acute kidney injury (AKI), only 50 out of 223 AKI patients required referral to the renal specialized hospital due to the persistence of acute kidney injury (AKI). Studies also describe that a portion of patients without renal replacement therapy (RRT) recovered spontaneously with optimal sepsis care [29-30]. The findings of our study support and greatly extend those of previous findings. Metabolic acidosis is also anticipated in severe sepsis. Severe sepsis can produce metabolic acidosis in a variety of ways. Either insufficient oxygen delivery with tissue hypoxia and resultant anaerobic glycolysis [31] or overproduction of pyruvate [32] may lead to lactic acidosis in severe sepsis. Septic acute kidney injury (AKI) makes the kidney unable to excrete waste products of nitrogen metabolism (urea and creatinine) which also contributes to acidosis [33]. Though recommended by SSC guidelines, in our settings, we could not check the serum lactate level of septic patients due to economic constraints.

Calcium in its active form (ionized Ca) can diffuse across cellular membranes to maintain body homeostasis [34]. Hypocalcemia, in association with severe sepsis [35], is commonly found in hospitalized critically ill patients [36]. In our study, we observed that hypocalcemic patients were at higher risk of having severe sepsis. Here, we have the facilities to measure total calcium and we used to treat hypocalcemic patients with intravenous infusion of calcium gluconate followed by oral supplementation of calcium carbonate. Contrasting evidence exist, about the role of calcium supplementation in severe sepsis. A Cochrane review concluded that supplementation has no added benefit, [37] whereas a recent retrospective study demonstrated calcium supplementation has a protective role for 28-days mortality [38].

### Study limitations

The main limitation of our study was that we were unable to perform a follow-up of patients who were referred for management of myocardial infarction and renal failure and thus their outcome was not known to us. This might have an impact on having only 9% mortality among our study population with severe sepsis, which is lower than those in other studies. On the other hand, we also have some strengths of our study. It was a five-year follow-up study involving a good number of patients. Dhaka hospital of icddr,b has a standard protocol that follows SSC guidelines. So, the quality of care did not differ much. Although retrospective observation, this is the first study where we have evaluated severe sepsis among adults who were hospitalized for diarrhea.

## Conclusion

Our data suggest that severe sepsis is common in diarrheal adults and the rate of progression from severe sepsis to septic shock is high. Cases had a higher case-fatality-rate than the controls. Systemic steroid intake, ileus, acute kidney injury (AKI), metabolic acidosis, and hypocalcemia were independently associated with severe sepsis in diarrheal adults. In resource-poor settings like ours, proper history taking, rigorous follow-ups, and early identification of organ dysfunction are essential for a better outcome.

## Data Availability

Data would be available from the Research administration, icddr,b when appropriate.

## Acknowledgements

We gratefully acknowledge the core donors for their support and commitment to icddr,b’s research efforts. We would like to express our sincere thanks to all clinical fellows, nurses, members of the feeding team, and cleaners of the hospital for their invaluable support and contribution to patient care.

